# ABC of Endometriosis Surgery: Aqua Blue Contrast Technique

**DOI:** 10.1101/2020.02.27.20027888

**Authors:** Tamer Seckin, Bahar Yuksel, Serin Seckin, Ayse Ayhan

**Affiliations:** Lenox Hill Hospital-Northwell Health Department of Obstetrics and Gynecology, New York; Endofoundation of America, Northwell Health Obstetrics and Gynecology Department, New York; Department of Gynecology, Mt. Sinai St. Luke’s and Mt. Sinai West, New York; Department of Pathology, Seirei Mikatahara Hospital, and the Department of Tumor Pathology, Hamamatsu University School of Medicine Hamamatsu, and the Department of Molecular Pathology, Hiroshima University School of Medicine, and the department of pathology

## Abstract

**Objective:** Peritoneal endometriosis is the most prevalent yet least diagnosed type due to its unrecognizable nature on imaging modalities especially non- pigmented lesions would escape the surgeons’ eye and experience during diagnostic laparoscopy. We used color adjuvant by the technique called Aqua Blue Contrast Technique (ABCT) to optimize the view and to improve results.

**Material methods:** Patient charts who undergone surgery from 2014 to 2015 and their 5 year follow up data, along with two different control groups, have been analyzed retrospectively. As the first group the patients who had conventional surgery without the use of ABCT) were included, the second group were patients who had ABCT applied on both pelvic side walls but not in cul-de-sac and as the third group the patients who had the surgery with the use of ABCT in all peritoneal cavity have been analyzed. Cases involving ovarian endometriomas and DIE were excluded.

**Results:** All patients have been followed up for up to 5 years. In group 1, the recurrence within the postoperative 3 years was 11.9%, and 16.1% after 5 years of surgery. The recurrence of 3 years period and 5 years period for the patients in group 2 were 8.7& and 12.1% respectively. The patients in group 3 recurred 3.2% in the 3 years period and the 4.3% recurrence rate calculated as 4.5% for the following 5 years after surgery.

**Conclusion:** Results indicate elimination of high-end spectrum of light using aqua blue contrast technique with hydrodistension of the retroperitoneum enhances the surgeon’s vision, allows lesions otherwise not detected with white light.

## Introduction

What is invisible to human eye does not mean its non-existence, and science always have its way to find it! This is one of the many situations where physics merges into medicine to solve problems.

Laparoscopy offered magnified access to the cavities, including the pelvic cavity eventually leading to recognition peritoneal endometriosis lesions otherwise not recognized during laparotomy. Late 1980’s and early 1990’s pigmented and non-pigmented occult lesions were described(1, 2). Since then no further discussions, and descriptions concerning the non-typical lesions were subject of any publication for about 30 years. A narrow band imaging via the couplers light source as well as color adjuvants such as ICG - fluorescent dye and indigo carmine were used to detect early angiogenesis of endometriosis lesions, with no clear documented diagnostic advantage(3-5). Here we offer a simple, cost effective novel technique to identify wide range of non-pigmented occult peritoneal lesions otherwise not recognized under the routine bright laparoscopic white light.

Endometriosis is an estrogen sensitive stem cell driven chronic inflammatory condition of the female pelvis characterized by the presence of endometrium like lesions outside of the uterine cavity(6, 7). Subsequent to visual recognition, the definitive diagnosis is based on surgical excision and histopathological examination of the resected tissue under the microscope for verifying endometrial glands and stroma. Based on surgically proven lesions that are excised, the prevalence of endometriosis in women on their reproductive ages all over the world, is at least 10% (2, 3). The real prevalence is still unknown because of underdiagnosis due to the lack of visibility/perceptibility hence to capture endometriotic lesions during surgery.

Endometriosis, as described by Nisolle and Donnez, is considered to have three anatomical subtypes, which are peritoneal endometriosis, ovarian endometriomas and deep infiltrative endometriosis (DIE)(8-10). Peritoneal endometriosis is the most prevalent yet least diagnosed type due to its unrecognizable nature on imaging modalities while ovarian endometriomas and deep infiltrative endometriosis lesions can be readily detected by ultrasound or MRI imaging (11). Furthermore, the preponderant presence of non-pigmented lesions would escape the surgeons’ eye and experience during diagnostic laparoscopy. In fact, most of the surgeries performed for deep endometriosis or ovarian endometriomas usually fail and may necessitate re-surgery upon insufficient excision of the peritoneal disease (12). Overall the lesions on the peritoneum can be scant in number, yet the presence of diffuse miliary and aggressive lesion is not uncommon. Therefore, defining this condition as “superficial” endometriosis may underscore the validation of multiple organ symptoms. Missing the lesions and so the disease cause misleading, misdiagnosing these patients as irritable bowel syndrome or pelvic floor disfunction hence undertreatment. Not only the problems of recurrence, occurrence of new lesions or the growth of occult ones, but also many symptoms including infertility, all should be based on the above facts (13)

The cardinal presentations of endometriosis are progressive pelvic pain primarily initiated with dysmenorrhea, gastrointestinal symptoms and infertility (14). Whether it can be thought the most devastating anatomical lesion may cause more pain, the pain of endometriosis is usually unrelated with the characteristics of pathology or severity of the disease (15). Therefore, a small lesion even not visible to the naked eye may cause more pain than a large DIE nodule. This knowledge further strengthens the importance of appropriately treating all lesions on a symptomatic patient with pain.

The standard surgical technique to diagnose and treat endometriosis is laparoscopic surgery. Under laparoscopic scope, endometriotic lesions can readily be identified when they are pigmented, also known as classical typical lesions as described by Sampson back in 1920’s(16). These lesions are in black, red, blue raspberry. Occult types of white colors and as flat or polypoid are described later 1n 1980’s (10). Although human eye can see wavelengths between 400-700 nm, it is not always easy to visualize all pathologies even with the 4 to 40 times magnification of usual laparoscopic scopes(17).Reflection of underlying red and yellow hues prevents discrimination of different tissue types under white light (18). As certain color combinations provide better contrast than others like, red and yellow against blue; to make objects easy to be caught by human eye using color contrasts would be an important advantage. Given the orange-red color of the intraabdominal tissues a blue dye would provide the best color contrast.

In our novel, Aqua Blue Contrast Technique™(ABCT) for endometriosis surgery we used methylene blue (methylene blue-methylthioninium chloride, C16H18N3SCl) for its natural blue color to make endometriotic lesions more visible and to filter red, yellow and white colors reflecting from peritoneal surface yet endometriosis primarily of its initial stage a peritoneal process. We aimed to evaluate the advantage of the technique to detect the peritoneal occult lesions, to see and compare the difference in endometriosis lesions, recurrence rates and quality of life changes within the same patient which ABC is used on pelvic side walls but not in cul-de-sac, also in patients groups that the technique is applied and not applied.

## Material Methods

After institutional review board (IRB) approval (17-0645-NH) has been taken, patient charts who undergone surgery from 2014 to 2015 and their 5 year follow up data, along with two different control groups, have been analyzed retrospectively. As the first group the patients who had conventional surgery without the use of ABC technique (ABCT) were included, the second group were patients who had ABCT applied on both pelvic side walls but not in cul-de-sac and as the third group the patients who had the surgery with the use of ABCT in all peritoneal cavity have been analyzed. All patients who had suspicious endometriosis according to the pain symptoms but has no DIE nodules or endometriomas either in transvaginal ultrasound or in Magnetic Resonance Imaging (MRI) techniques and histologically proven peritoneal endometriosis after surgical removal have been included into the study. Cases involving ovarian endometriomas and DIE were excluded. All patients gave informed consent that their surgical notes can be used on a scientific research manner.

In the second and third groups a novel surgical approach has been used which includes a blue dye contrast usage, a unique aspect of excision surgery procedure called Aqua Blue Contrast Technique™(19). A solution made of 1 % methylene blue in 3000 cc isotonic sodium chloride has been prepared for the use at the surgery. All surgeries were done by the same trained endoscopic surgeon (TS). Laparoscopic approach using 10 mm umbilical trocars and 2 or 3 5 mm sister trocars has been placed. After visual inspection of pelvic and peritoneal organs, first phase involve hydro floatation, and submersion. MB is flushed, filled to the pelvic peritoneal cavity catch vision of floating or vegetative lesions under blue liquid which we called as immersion phase. And after sucking out all blue water, the second step which is called as retroperitoneal hydrodistension phase was initiated with a small 0.5cm excision was made on the peritoneum medial to IP ligament at the pelvic brim. Through the incision, laparoscopic irrigation tip which is connected to the ABC dye, was advanced into retroperitoneum of the pelvic sidewalls. Under 200 mmHg irrigation pressure retro peritoneal connective tissue space was hydro distended with diluted blue dye adjuvant Methylene Blue (20). All peritoneal cavity were inspected under direct submersion, but ABC hydro distention was only performed on the pelvic sidewalls while cul-de-sac has left for bare eye recognition in group 2 and all retroperitoneum was hydro distended in group 3. All suspected lesions for endometriosis have been excised by cold scissors and specimens send for histopathological investigation. No adverse effects related to the MB use have been reported. Hematoxylin and eosin (H&E) staining has been used for microscopic evaluation and diagnosis. And all specimens have been undergone histopathological evaluation and classified according to the results.

After all cases were collected their files were searched for following 5 years period and patients who are re-operated because of pain recurrence have been noted.

Statistical analyses were performed using the version 20.0 of Statistical Package for the Social Sciences software (SPSS, Inc, Chicago, Illinois). A P value of <0.05 was considered significant.

## Results

The pathology reports were analyzed, and histological diagnosis were noted as typical endometriosis, stromal endometriosis without glands in exhausted serial sections, inflammation, and fibrosis, documented in Table …In group one 371 cases have been reviewed. Out of 1452 samples taken from all locations 1383(95.2%) were found to be typical endometriosis, 13(0.95%) as stromal endometriosis, 56 (3.9%) as inflammation and/or fibrosis. In group 2 out of 775 samples, 407 were found to be typical endometriosis, 49 stromal endometriosis, 237 inflammation and 82 fibrosis. Six hundred sixty-three samples were collected from pelvic side walls where ABC technique have been applied but only 112 suspected lesions have been detected without blue contrast from cul-de-sac (z=2.133, p<0.05) The distribution of pathology diagnoses of the samples taken from cul-de-sac were; 58 (52%) typical endometriosis, 2 (2%) samples for stromal endometriosis, 52 (47%) inflammation and/or fibrosis, while 663 samples from pelvic side walls came out as 349 (53%) typical endometriosis, 47 stromal endometriosis (7%), 267 (40%) inflammation and or fibrosis. The evaluation of the pathological diagnoses regarding typical endometriosis, fibrosis and inflammation showed similar distribution while the proportion of stromal endometriosis significantly greater in pelvic sidewall excisions compared to cul-de-sac excisions (47 sidewalls, 2 cul de sac; z=19.79, p<0.0001). And out of 7080 samples taken from 684 patients in group 3, 5217 (73.7%) samples analyzed as typical endometriosis, 476 (6.7%) as stromal endometriosis, 1387 (19.5%) as inflammation and/or fibrosis.

All patients have been followed up for up to 5 years. In group 1, the recurrence within the postoperative 3 years was 11.9%, and 16.1% after 5 years of surgery. The recurrence of 3 years period and 5 years period for the patients in group 2 were 8.7& and 12.1% respectively. The patients in group 3 recurred 3.2% in the 3 years period and the 4.3% recurrence rate calculated as 4.5% for the following 5 years after surgery. When the histopathological evaluation of surgically resected material from recurrent 14 patients in group 2, where the ABCT partially used, were analyzed, 25 pelvic side wall [13 (52%) typical endometriosis, 9(36%) inflammation and 3(12%) fibrosis] and 84 cul-de-sac [53 (63%) typical endometriosis, 5(6%) inflammation, 26(31%) fibrosis]. The statistical comparison of the number and distribution of histopathology results between pelvic side wall and cul-de-sac samples did not reveal any statistical significance (p=0.60). In the recurrent operation, additionally two patients needed DIE nodule resection, 8 patients required endometrioma surgery, 1 patients had endometrial polyp resection, 3 patients had appendix removal because of being affected by endometriosis.

## Discussion

MB by having minimal and benign side effects as dizziness, head ache, diarrhea and also very rarely excitation caused by its monoamine oxidase inhibitor (MAOI)function, found its place in World Health Organization’s essential medicine medications list(21).

ABC technique provides a laparoscopic surgery method allowing better inspection of peritoneal surfaces under a blue colored solution, by a contrast enhancing agent, typically using near contact scanning by a laparoscope. This technique easily identified normal peritoneum and its texture and successfully distinguished it from abnormal peritoneal surfaces. This method assists the surgeon to precisely target the lesion by distinct recognition of even most subtle lesions and associated peritoneal changes in endometriosis. Eliminating the yellow and red hues by blue retroperitoneal contrast, it helps to perform tedious, precise and accurate excision surgery without indiscriminate removal of normal peritoneum(22).

This study however showed that by the aid of ABC technique, it is possible to recognize and remove more endometriosis lesions from pelvic sidewalls which was assumed to be less effected compared to cul-de-sac and also from all over the peritoneal area that the technique has been applied Also long term results showed that using ABCT prevented recurrence of the disease significantly.

Changing the color spectrum and using hydro floatation with contrast color and retroperitoneal distention may help to visualize the morphological features of the peritoneum along with endometriosis otherwise undetectable by standard laparoscopic light inspection. Further, retroperitoneum hydro-distention and creation of blue color contrast in the background makes it possible to identify peritoneal micro defects to determine the excision boundaries and allows inspection of retroperitoneal space under blue colored solution for microinvasion and fibrosis (23). Under these circumstances, the blue color infused into the connective tissue of retroperitoneal organs and peritoneum itself acts as a blue color filter just reflecting only blue light and eliminating the interfering all reds and yellows. By absorbing and blocking red and green colors, blue reflection in addition contributes visual representation of only blue light, therefore objects with contrast color light component would seem without a color which is black to human eye(24). Red is a primary color while yellow on one perception a secondary color and, made of green and red light so this explains any kind of red or yellowish spot would appear as black spot under blue filter(25) On the other hand, according to Isaac Newton’s almost 300 years old color wheel theory, primary colors are three: blue, red and yellow. Red and yellow represent the opposite spectrum, meaning they are contrast colors of blue (26). Also, fibrotic tissues more distintive features, conspicuously recognized with white, and off-white nature towards blue effluence. This might explain why all endometriotic lesions seem more visible to human eye under Aqua Blue Contrast Technique™.

According to the retrograde menstruation theory of Sampson’s, as also quoted by published works of others, cul-de-sac is the most common location for peritoneal endometriosis (27). However, a trained endometriosis surgeon should be aware that the non-pigmented endometriosis lesions constitute the majority of the disease and are unrecognizable by human eye because of the color and light reflections under white light of the scope.

The easy-to-visualize types of endometriosis, namely the pigmented lesions, are typically red, purple or black, and easily recognized over others which are subtle and invisible, also called as microscopic endometriotic lesions (1, 28, 29). The results of our study may not be enough to say that the pelvic side walls are affected more than cul-de-sac, but definitely mean to say that the color contrast technique efficiently helps human eye to recognize distorted areas easily. In the present study, we reached at the conclusion that aided by Aqua Blue Contrast Technique™, the excision rate multiplied by at least 2.13 times compared to unaided detection under laparoscopic light.

Laparoscopic surgery is the only standard availing both diagnosis and treatment of endometriosis. The efficiency of the primary surgery increases patient’s quality of life whereas incomplete excision risks symptom relapses and recurrent surgeries (15, 26). In our study the five year follow up of the patients with focal excision of endometriosis lesions using Aqua Blue Contrast Technique™ showed a recurrence rate resulting in 13% reoperation, while studies in the literature showed more than 30% recurrence after 36 months follow up of the patients who are operated by conventional techniques (30).

Pelvic pain with dysmenorrhea is the cardinal symptom of endometriosis but the underlying mechanism is remains inconclusive(31). The studies aiming to specify a correlation between lesion feature and pain symptoms have been unsuccessful. It is not known whether the pain is caused by endometriosis itself or due to the fibrosis or inflammation (32). However numerous studies demonstrated that the angiogenesis component of endometriosis also reveal neurogenesis along with the macrophage activity in the developing new lesions(33) The resection of all lesions along with fibrotic distortions caused by endometriosis with the aim of restoring anatomy, remains critical center core of the treatment. Previous studies showing endometriosis as a stem cell derived estrogen sensitive metaplastic inflammation with the presence of gland and/or stroma increases the importance of resection of all effected tissues(7). The effect of fibrosis, inflammation and stromal endometriosis should be investigated in further studies.

The limitations of this study is its retrospective character, however, has enough power when the patient numbers are concerned. The in vivo effect, the chance to compare lesions on each same patient and all operations being done by the same surgeon give further strength to the study. Also, the low cost, simple applicability of the technique over other color adjuvants or narrow-band light techniques, and the safety without any side effect, offer more opportunity along with a wide range utilization.

## Conclusion

Identification of the disease by the surgeons is only possible by recognizing the lesion based on its visual characteristics. Failing the visual recognition of different endometriosis lesions and missing the non-colored lesions would lead to incomplete surgery, lead to inaccurate data negatively impacting forthcoming research and scholarly reviews, and most importantly leading to treatment failure.

Results indicate elimination of high-end spectrum of light using aqua blue contrast technique with hydrodistension of the retroperitoneum enhances the surgeon’s vision, allows lesions otherwise not detected with white light. We have shown the evidence by increased number of excision specimens and increased proportion of stroma-positive specimens with Aqua Blue Contrast Technique™. It is notable that statically significant increase of cul-de-sac lesions in repeat surgeries would imply either the persistence of the disease possibly missed due to not using Aqua Blue Contrast Technique™, or de novo appearance of new lesions in favor cul-de-sac over previously excised pelvic sidewalls.

**Table 1:** Distribution of pathology results

| Patients (n) |  |  |  |  |  |
| --- | --- | --- | --- | --- | --- |
| samples (n) |  | Endometriosis |  |  |  |
| n (%) | Stromal endometriosis |  |  |  |  |
| n (%) | Inflammation+/- fibrosis |  |  |  |  |
| n (%) | P value |  |  |  |  |
| Group 1 | 371 | 1452 | 1383 (95.2%) * | 13(0.95%) | 56(3.9%) |
| Group 2, CDS | 115 | 112 | 58 (52%) | 2(2%) | 52(47%) |
| Group 2, PSW | 115 | 663 | 349(53%) | 47(7%) | 267(40%) |
| Group 3 | 684 | 7080 | 5217(73.7%) * | 476(6.7%) | 1387(19.5%) |
| P value | P<0.001 |  |  |  |  |

**Table 2:**
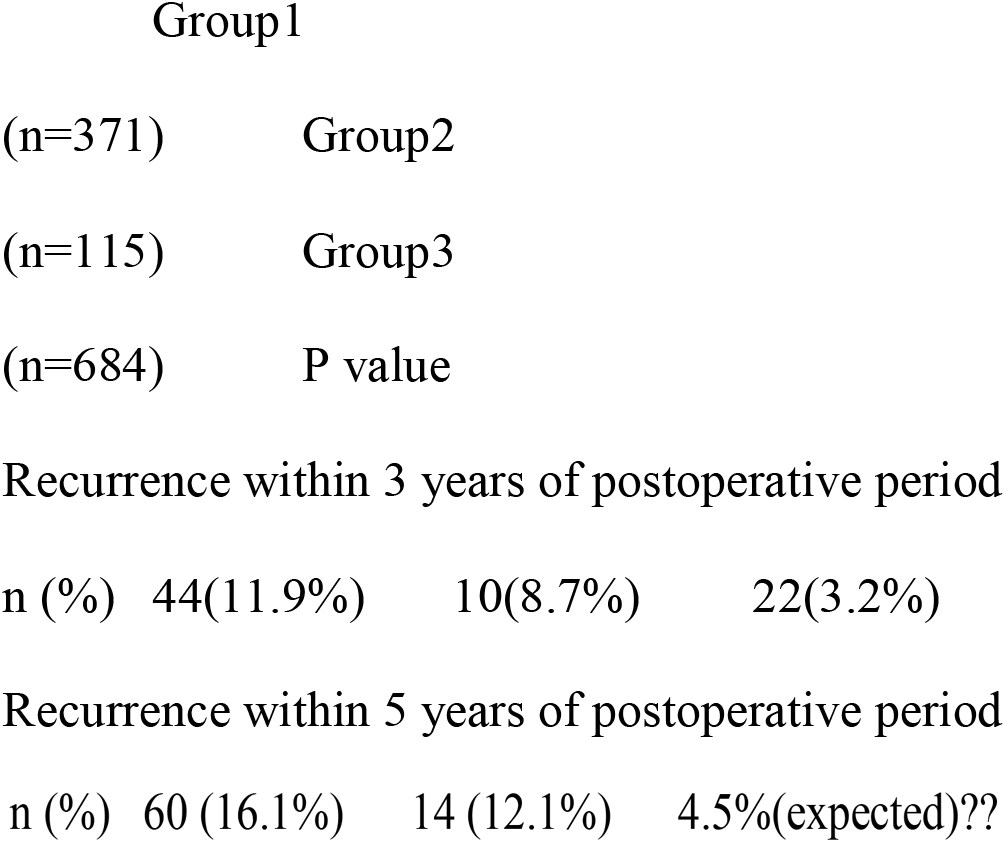
Recurrence ratios of each groups for 3 and 5 years follow up periods

**Figure 3:**
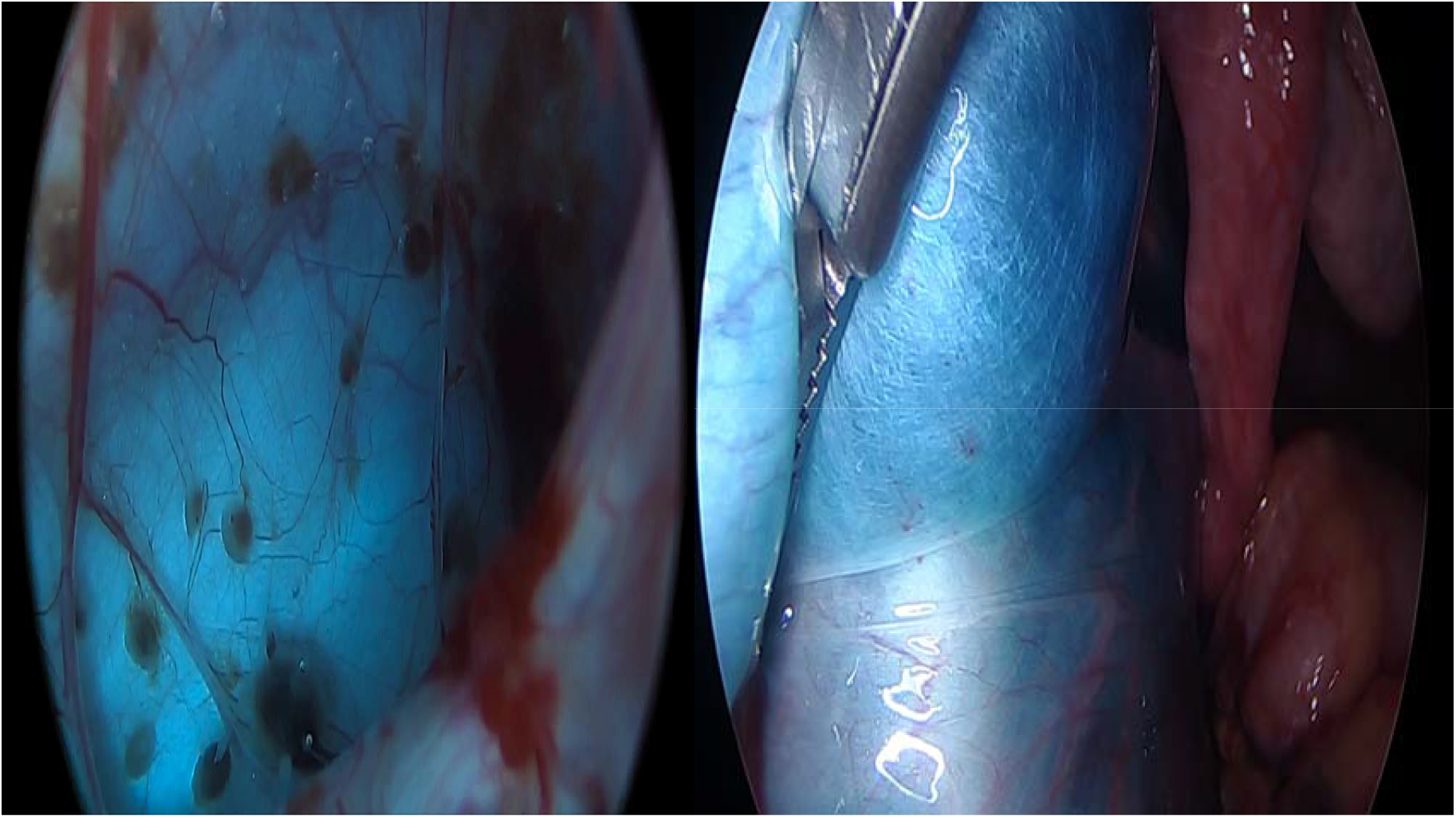
Picture of normal healthy peritoneum under ABC technique and peritoneum effected by endometriosis Isaac Newton - Optics, 4th ed., 1730. From Book I, Part II, Proposition VI, Problem 2. This figure is nearly unchanged from that in the 1704 first-edition printing. The primary and secondary colors on light and color palette spectrum (from right to left)

## Data Availability

data is available in case of requested

## References

1. Redwine DB. Age-related evolution in color appearance of endometriosis. Fertility and sterility. 1987;48(6):1062–3.

2. Martin DC, Hubert GD, Vander Zwaag R, el-Zeky FA. Laparoscopic appearances of peritoneal endometriosis. Fertility and sterility. 1989;51(1):63–7.

3. Rauh-Hain JA, Laufer MR. Increased diagnostic accuracy of laparoscopy in endometriosis using indigo carmine: a new technique. Fertility and sterility. 2011;95(3):1113–4.

4. Cosentino F, Vizzielli G, Turco LC, Fagotti A, Cianci S, Vargiu V, et al. Near-infrared imaging with indocyanine green for detection of endometriosis lesions (Gre-Endo Trial): a pilot study. Journal of minimally invasive gynecology. 2018;25(7):1249–54.

5. Al-Taher M, Hsien S, Schols RM, Hanegem NV, Bouvy ND, Dunselman GAJ, et al. Intraoperative enhanced imaging for detection of endometriosis: A systematic review of the literature. European journal of obstetrics, gynecology, and reproductive biology. 2018;224:108–16.

6. Alimi Y, Iwanaga J, Loukas M, Tubbs RS. The Clinical Anatomy of Endometriosis: A Review. Cureus. 2018.

7. Anglesio MS, Papadopoulos N, Ayhan A, Nazeran TM, Noë M, Horlings HM, et al. Cancer-Associated Mutations in Endometriosis without Cancer. The New England journal of medicine. 2017;376(19):1835–48.

8. Bulun SE, Yilmaz BD, Sison C, Miyazaki K, Bernardi L, Liu S, et al. Endometriosis. Endocrine reviews. 2019;40(4):1048–79.

9. Nisolle M, Donnez J. Peritoneal endometriosis, ovarian endometriosis, and adenomyotic nodules of the rectovaginal septum are three different entities. Fertility and sterility. 1997;68(4):585–96.

10. Koninckx PR, Ussia A, Adamyan L, Wattiez A, Gomel V, Martin DC. Heterogeneity of endometriosis lesions requires individualisation of diagnosis and treatment and a different approach to research and evidence based medicine. Facts, views & vision in ObGyn. 2019;11(1):57–61.

11. Noventa M, Scioscia M, Schincariol M, Cavallin F, Pontrelli G, Virgilio B, et al. Imaging Modalities for Diagnosis of Deep Pelvic Endometriosis: Comparison between Trans-Vaginal Sonography, Rectal Endoscopy Sonography and Magnetic Resonance Imaging. A Head-to-Head Meta-Analysis. Diagnostics (Basel, Switzerland). 2019;9(4).

12. Jayot A, Bendifallah S, Abo C, Arfi A, Owen C, Darai E. Feasibility, Complications, and Recurrence after Discoid Resection for Colorectal Endometriosis: A Series of 93 Cases. Journal of minimally invasive gynecology. 2020;27(1):212–9.

13. Reis FM, Santulli P, Marcellin L, Borghese B, Lafay-Pillet M-C, Chapron C. Superficial Peritoneal Endometriosis: Clinical Characteristics of 203 Confirmed Cases and 1292 Endometriosis-Free Controls. Reproductive sciences (Thousand Oaks, Calif). 2020:10.1007/s43032-019-00028-1.

14. Martin DC, Ling FW. Endometriosis and pain. Clinical obstetrics and gynecology. 1999;42(3):664–86.

15. Fauconnier A, Chapron C, Dubuisson JB, Vieira M, Dousset B, Breart G. Relation between pain symptoms and the anatomic location of deep infiltrating endometriosis. Fertil Steril. 2002;78(4):719–26.

16. Sampson JA. Metastatic or Embolic Endometriosis, due to the Menstrual Dissemination of Endometrial Tissue into the Venous Circulation. The American journal of pathology. 1927;3(2):93-110.43.

17. Yun SH, Kwok SJJ. Light in diagnosis, therapy and surgery. Nature Biomedical Engineering. 2017;1(1).

18. Hammond BR, Sreenivasan V, Suryakumar R. The Effects of Blue Light-Filtering Intraocular Lenses on the Protection and Function of the Visual System. Clinical ophthalmology (Auckland, NZ). 2019;13:2427–38.

19. Seckin TA, Newman NC, Seckin S. Histological Findings Following Excision of Peritoneal Endometriosis With and Without Using Aqua Blue Contrast Technique (ABCt™). Journal of Minimally Invasive Gynecology. 2015;22(6):S52.

20. Tardivo JP, Del Giglio A, de Oliveira CS, Gabrielli DS, Junqueira HC, Tada DB, et al. Methylene blue in photodynamic therapy: From basic mechanisms to clinical applications. Photodiagnosis and Photodynamic Therapy. 2005;2(3):175–91.

21. Tucker D, Lu Y, Zhang Q. From Mitochondrial Function to Neuroprotection-an Emerging Role for Methylene Blue. Mol Neurobiol. 2018;55(6):5137–53.

22. Sturmey RG, Wild CP, Hardie LJ. Removal of red light minimizes methylene blue-stimulated DNA damage in oesophageal cells: implications for chromoendoscopy. Mutagenesis. 2009;24(3):253–8.

23. Lessey BA, Higdon HL, Miller SE, Price TA. Intraoperative Detection of Subtle Endometriosis: A Novel Paradigm for Detection and Treatment of Pelvic Pain Associated with the Loss of Peritoneal Integrity. Journal of Visualized Experiments. 2012(70).

24. Conway BR. Color vision, cones, and color-coding in the cortex. Neuroscientist. 2009;15(3):274–90.

25. Gismondi E. Polychrome lighting device having primary colors and white-light sources with microprocessor adjustment means and remote control. Google Patents; 1999.

26. Newton IOp. Optics PP 114–117. 1704.

27. Sampson JA. The development of the implantation theory for the origin of peritoneal endometriosis. American Journal of Obstetrics and Gynecology. 1940;40(4):549–57.

28. Donnez J, Van Langendonckt A. Typical and subtle atypical presentations of endometriosis. Curr Opin Obstet Gynecol. 2004;16(5):431–7.

29. Redwine DB. ‘Invisible’ microscopic endometriosis: a review. Gynecologic and obstetric investigation. 2003;55(2):63–7.

30. Vercellini P, Aimi G, Busacca M, Apolone G, Uglietti A, Crosignani PG. Laparoscopic uterosacral ligament resection for dysmenorrhea associated with endometriosis: results of a randomized, controlled trial. Fertility and sterility. 2003;80(2):310–9.

31. Tokushige N, Markham R, Russell P, Fraser IS. Nerve fibres in peritoneal endometriosis. Human Reproduction. 2006;21(11):3001–7.

32. Morotti M, Vincent K, Becker CM. Mechanisms of pain in endometriosis. European journal of obstetrics, gynecology, and reproductive biology. 2017;209:8–13.

33. Asante A, Taylor RN. Endometriosis: the role of neuroangiogenesis. Annu Rev Physiol. 2011;73:163–82.

